# Predicting Disease Progression in Critically Ill Patients Using Frequency-Enhanced Time-series Forecasting

**DOI:** 10.64898/2025.12.05.25341734

**Authors:** Zhengxu Li, Chunyu Hu, Wei Zhuang, Zengjie Dong, Hong Liu, Wenhao Li

## Abstract

**Purpose:** Accurate disease progression prediction is vital for managing critically ill patients in intensive care. Existing deep learning approaches mainly operate in the time domain and often fail to capture long-range dependencies and spectral dynamics. This study proposes a unified framework integrating time and frequency-domain representations to improve predictive accuracy.

**Methods:** We introduce FETT (Frequency-Enhanced TCN-Transformer), a dual-domain forecasting framework that combines discrete wavelet transform–based frequency analysis with transformer-based temporal modeling. Based on an iTransformer backbone, FETT introduces three key innovations: (1) a frequency-aware representation module, (2) a dual-TCN architecture that enhances temporal representation through multi-scale feature extraction and global dependency modeling; and (3) a frequency-aware inverse reconstruction module for clinically interpretable time-domain forecasts.

**Results:** Experiments on the MIMIC-IV Sepsis-3 cohort show that FETT outperforms state-of-the-art baselines, reducing MSE by up to 20.81% and MAE by 13.55% in 24-hour forecasting tasks. Ablation studies confirmed the complementary contributions of the dual-TCN design and frequency-aware modules.

**Conclusion:** FETT effectively integrates time- and frequency-domain information to deliver accurate and interpretable disease progression predictions in critical care. By bridging spectral and temporal representations, it enables early detection of patient deterioration and holds strong potential for advancing proactive ICU monitoring and personalized clinical decision-making.

## 1. Introduction

Accurate disease progression prediction is vital for managing critically ill patients, whose conditions are often highly dynamic and rapidly evolving. In critical care settings, such as intensive care units (ICUs), clinicians must closely monitor several key physiological signals, such as heart rate, blood pressure, respiratory rate and oxygen saturation, to identify early signs of physiological instability. When combined with advanced predictive capability that proactively assesses patient trajectories, such continuous physiological monitoring enables early risk stratification and guides timely, evidence-based clinical interventions, thereby enhancing patient outcomes and mitigating the risk of deterioration [1].

Physiological signals, with their temporal dynamics, complexity, and multivariate nature, are key to disease progression prediction. By leveraging historical physiological signals, machine learning models can automatically detect patterns indicative of impending clinical deterioration and generate timely predictions to assist in decision-making. Among existing machine learning approaches, deep learning models, including recurrent neural networks (RNNs), long short term memory (LSTM) networks, and transformer architectures, are effective for modeling temporal dependencies and complex nonlinear patterns in physiological signals [2–4].

Traditional methods of predicting disease progression have primarily framed the problem as binary classification or regression. These methods produce single-valued outputs that assess specific outcomes (e.g., mortality or ICU admission rate) or variations in individual physiological signals [5–8]. These methods offer limited assistance for informed clinical decision-making, as they fail to account for the temporal continuity and interdependence of physiological variables that characterize disease evolution [9]. Recently, growing emphasis has been placed on time-series forecasting (TSF) tasks to capture the temporal evolution of physiological signals [10]. These tasks leverage patients’ historical physiological signals to predict future values. This paradigm enables the continuous monitoring of multiple parameters over extended periods, providing forward-looking estimates that allow clinicians to anticipate patient deterioration and intervene at an earlier stage. However, existing TSF models for disease progression prediction are constrained by two key limitations. First, existing models often fail to consider the frequency-domain features inherent in physiological signals, thereby limiting their ability to fully exploit periodic patterns that characterize cyclic physiological processes and multi-scale dependencies [11]. Without dedicated frequency-domain analysis, these models struggle to detect periodic anomalies and intrinsic fluctuations in vital signs (e.g., heart rate and respiration). This creates an information bottleneck that degrades predictive performance. Second, existing methods mostly rely on point-wise mapping strategies, i.e., they generate outputs based on local correlations at the current or adjacent time steps, without incorporating multi-scale modeling to capture long-range temporal dependencies across the entire sequence [12]. For example, recurrent and convolutional models often rely on local temporal correlations within limited receptive fields, restricting their ability to capture long-range dependencies and global temporal dynamics [13]. Consequently, these strategies fail to capture global temporal patterns (e.g., hour-to-day trends in patient status) that are essential for understanding signal trajectories and disease progression.

To address these limitations, we propose **FETT (Frequency-Enhanced TCN-Transformer)**, a unified framework that integrates temporal and frequency-domain representations to capture both long-range dependencies and periodic patterns in physiological signals. First, to address the lack of frequency-domain modeling, FETT introduces the extraction of frequency-domain features from physiological signals and learns frequency- and time-domain representations separately. Specifically, the frequency-domain stream decomposes vital sign signals to extract their periodic features and potential anomalies [11]. Working in parallel with the time-domain stream, it integrates frequency-domain representations with laboratory measurements and demographic attributes to facilitate joint training and holistic representation learning. Second, to move beyond point-wise mappings and better capture global temporal dependencies, FETT builds upon the iTransformer architecture by integrating a complementary dual-TCN design [14] that enhances temporal representation learning and improves optimization stability. Specifically, in the embedding phase, one TCN extracts multi-scale temporal features to produce representations with explicit temporal ordering, thereby introducing temporal structural awareness into the framework. In the encoder, a second TCN models dynamic interactions across time steps to effectively capture long-range dependencies, further extending the temporal perception of the iTransformer and facilitating the fusion of global and local information within a unified encoding layer. Lastly, to produce human-readable output that can be directly interpreted by clinicians, FETT merges frequency-domain features back into the time domain to generate the final output—a prediction of physiological signals over a specified future period. The predictive output captures deviations of physiological signals from baseline trajectories, allowing clinicians to anticipate potential physiological deterioration, thereby improving the effectiveness of real-time ICU monitoring and individualized interventions, and hence enhancing final treatment outcomes.

The main contributions of this paper are as follows:

- It proposes a physiological signals-based disease progression prediction framework that integrates both temporal and frequency-domain information to enhance prediction accuracy, which generates future physiological signals that can be directly interpreted by clinicians.
- It introduces an innovative dual TCN design into the iTransformer backbone to strengthen the framework’s awareness of temporal structures, and enhance its ability to capture both short- and long-term temporal dependencies, thereby improving overall time-series forecasting performance.
- Extensive experiments based on MIMIC-IV sepsis cohorts data were conducted. Results show that FETT consistently outperforms state-of-the-art baseline models across multiple evaluation metrics including Mean Squared Error (MSE) and Mean Absolute Error (MAE). Additional ablation studies further validate the effectiveness of the frequency-aware modules and the contribution of the dual TCN design.

The remainder of this paper is organized as follows. “Related work” presents the related research on time series forecasting. “Experiments” presents the experimental setup. “Results and discussion” presents and discusses the results, and “Conclusion” concludes the paper and outlines future research avenues and potential improvements.

## 2. Related work

### 2.1 Deep learning for time-series forecasting

Deep learning–based approaches have emerged as powerful tools for time-series forecasting. Leveraging hierarchical representation learning, these approaches leverage hierarchical representation learning to capture nonlinear temporal patterns from historical time-series data to predict future trends. Early research has shown that Recurrent Neural Networks (RNNs) and their variants including Long Short-Term Memory (LSTM) and Gated Recurrent Unit (GRU) models, are effective in capturing temporal dependencies and nonlinear dynamics in time-series forecasting tasks [13, 15–17]. Recent efforts have refined architectures to improve predictive accuracy. For example, stacking multiple LSTM layers has been shown to enhance performance in univariate time-series forecasting tasks [18]. Beyond RNN-based designs, Transformer architectures leveraging self-attention mechanisms and parallel processing capabilities have substantially advanced sequence modeling and are now widely applied in time-series forecasting [19–23]. In [20], an inverted Transformer-based model named iTransformer was proposed, which independently embed each time-series variable, enabling more effective representation learning for multivariate forecasting.

### 2.2 Time-series forecasting in the medical field

Medical time-series data, such as physiological signals and laboratory measurements, contain rich temporal information that supports clinical decision-making. Leveraging this information, time-series forecasting has become essential in the medical field, enabling the anticipation of critical events and the tracking of disease progression [19, 24–26]. In [10], it introduced a Transformer-based model that uses continuous monitoring data from ICU patients to predict ongoing treatment outcomes, demonstrating superior predictive performance compared to traditional methods. In [19], it introduced the IS-LSTM model for blood glucose prediction in Type 1 diabetes patients. Using incremental retraining and parameter transfer, it learns individual glucose patterns from continuous glucose monitor data, thereby significantly reducing root mean square error compared to CNN-LSTM [27]. These approaches operate only in the time domain. Although spectral priors are used in some of these approaches (e.g., implicit frequency mixing), the frequency cues remain non-invertible, making their clinical meaning unclear and thereby limiting interpretability from a medical perspective.

### 2.3 Frequency-domain analysis of time-series data

Feature extraction from a frequency-domain perspective is becoming increasingly valuable for time-series forecasting tasks. Using time–frequency transformation techniques to convert signals from the time domain to the frequency domain enables models to exploit periodicity, spectral patterns, and latent temporal dynamics, and hence achieve a more comprehensive understanding of complex temporal behaviors and improved forecasting accuracy [28–31]. Classical Fourier-based transformation approaches provide stable global spectral features. They have been successfully applied to tasks such as EEG seizure classification, where frequency representations help distinguish seizures from artifacts [30]. Beyond Fourier analysis, wavelet-based methods have been widely adopted for capturing both time-localized and frequency-resolved features. In particular, the Discrete Wavelet Transform (DWT) with compactly supported wavelets (e.g., Daubechies-4) separates signals into low-frequency trends and high-frequency fluctuations, thereby enabling multi-scale feature extraction and enhancing the detection of transient events and subtle variations in the signal. In [32], it proposes a NICU neonatal sleep state stratification model, innovatively integrating multi-perspective feature extraction with Stationary Wavelet Transform (SWT), Flexible Analytic Wavelet Transform (FAWT), and spectral-temporal features, combined with Adaptive Feature Selection (AFS) and an ensemble stacking classifier (Random Forest, Extra Trees, XGB). It achieves over 85% accuracy on eight-channel EEG, significantly outperforming a range of base-line deep learning prediction models, demonstrating superior performance in neonatal sleep classification.

In this paper, the proposed FETT framework introduces several distinctive features that set it apart from previous works. Unlike FEDformer [22], which implicitly mixes frequency- and time-domain cues within the attention mechanism, FETT explicitly separates the frequency-domain stream from the time-domain process through an inverse reconstruction path. This design enables the frequency-domain contributions to be projected back into time-domain trajectories, thereby improving clinical interpretability while enhancing predictive accuracy. Compared with TimesNet [33], which models multivariate series through 2D variations, FETT adopts a variable-as-token formulation and incorporates two TCNs at different stages of the architecture: one in the embedding phase to capture ordinal semantics, and another in the encoder phase to model multi-scale temporal dependencies. Furthermore, unlike the baseline iTransformer [20], which employs attention followed by a position-wise feedforward neural network (FFN), FETT replaces the FFN with an encoder-TCN, introduces an embedding-TCN, and adds an explicit time–frequency stream with invertible back-projection. In summary, FETT complements global attention with explicit order encoding and multi-scale convolution, effectively bridging frequency-domain cues to clinically interpretable time-domain predictions, thereby facilitating practical bedside applications.

## 3. Methodology

This section provides a detailed description of the proposed FETT framework. It begins by formalizing the problem and defining the modeling objectives, then presents an overview of the overall architecture. The following subsections describe the design of each core module in detail, with particular emphasis on dual-domain feature extraction and temporal representation learning components.

### 3.1 Problem formulation

Vital signs and laboratory measurements are two essential types of physiological signals collected in a typical critical care scenario. Vital signs, including heart rate, blood pressure, respiratory rate, and body temperature, are critical indicators of basic physiological functions. Bedside monitoring systems continuously collect these signals, yielding high-resolution, real-time physiological time series. Meanwhile, laboratory measurements such as complete blood counts and urinalysis, are acquired using standardized protocols. These measurements are sampled at fixed intervals, producing discrete time series that encompass both quantitative and qualitative indicators of a patient’s physiological, pathological, and metabolic status. Given a patient’s demographic profile and vital sign and laboratory time series, the objective is to develop an automated forecasting framework capable of predicting future physiological trajectories covering certain time horizon. As shown in Table 1, which provides the definitions of the formula symbols.

**Table 1.** Summary of mathematical notations and variable definitions used.

| Symbol | Meaning | Range/Dimension |
| --- | --- | --- |
| T | Input step length | Integer (hours) |
| E | Prediction step length | Integer (hours) |
| N | Number of vital signs | Integer |
| M | Number of laboratory measurements | Integer |
| J | Number of demographic profiles | Integer |
| X | Historical time-series data | $\mathbb{R}^{(N+M+J) \times T}$ |
| $X_{fre}$ | Frequency-Enhanced Representation | $\mathbb{R}^{(2N+M+J) \times \lceil T/2 \rceil}$ |
| $\hat{Y}_{fre}$ | Frequency-Enhanced Forecast | $\mathbb{R}^{(2N+M) \times \lceil E/2 \rceil}$ |
| $\hat{Y}$ | Forecast time-series data | $\mathbb{R}^{(N+M) \times E}$ |

Formally,

#### Vital signs

Vital signs are approximated as discretely sampled time-series data to align with laboratory measurements. Let *S*_*n,t*_ denote the time-series data of the *n*-th vital sign, where *n* ∈ {0,1, …, *N* − 1} represents the type of the vital sign, and *t* ∈ {0,1, …, *T* − 1} indicates the discrete time step. The vector *S*_*t*_ = [*S*_0,*t*_, *S*_1,*t*_, …, *S*_*N*−1,*t*_]^⊺^ represents the collection of all vital signs collected at time step *t*, whereas *S*_*n*_ = [*S*_*n*,0_, *S*_*n*,1_, …, *S*_*n,T*−1_] denotes the complete time series corresponding to the *n*-th indicator over the monitoring period.

#### Laboratory measurements

Let *L*_*m,t*_ denote the time-series data of the *m*-th laboratory measurement, where *m* ∈ {0,1, …, *M* − 1} represents the type of the laboratory measurements, and *t* ∈ {0,1, …, *T* − 1} indicates the discrete time step. The vector *L*_*t*_ = [*L*_0,*t*_, *L*_1,*t*_, …, *L*_*M*−1,*t*_]^⊺^ represents the collection of all laboratory measurements at time step *t*, whereas *L*_*m*_ = [*L*_*m*,0_, *L*_*m*,1_, …, *L*_*m,T*−1_] denotes the complete time series corresponding to the *m*-th indicator.

#### Demographic profiles

Let *P*_*j*_ denote the *j*-th demographic profile attribute of a patient, where *j* ∈ {0,1, …, *J* − 1}. Each *P*_*j*_ is a scalar representing a time-invariant attribute (e.g., age at admission, sex, ethnicity). The collection of all demographic profile attributes for a single patient is denoted as *P*. In this study, demographic profile attributes are treated as static covariates. To ensure temporal alignment with the time-series data for subsequent analysis, the demographic profile *P* is broadcast along the temporal axis.

Let *X*_:,*t*_ = (*S*_*t*_, *L*_*t*_, *P*) denote the set of all physiological signals of a patient at time step *t*. The objective is to predict the physiological signals of subsequent *E* time steps 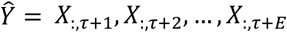 based on the physiological signals of previous *T* time steps *X* = [*X*_:,τ − *T*+1_, *X*_:,τ − *T*+2_, …, *X*_:,τ_] ∈ ℝ^*C*×*T*^, where *X* is a multivariate time series with *C* = *N* + *M* + *J* variables.

### 3.2 FETT overview

Figure 1 provides an overview of the FETT framework. From bottom to top, it consists of a Frequency Aware Representation module, an Augmented-Embedding module, an Encoder module, a Projection module, and a Frequency-Aware Inverse Representation module.

**Fig. 1.**
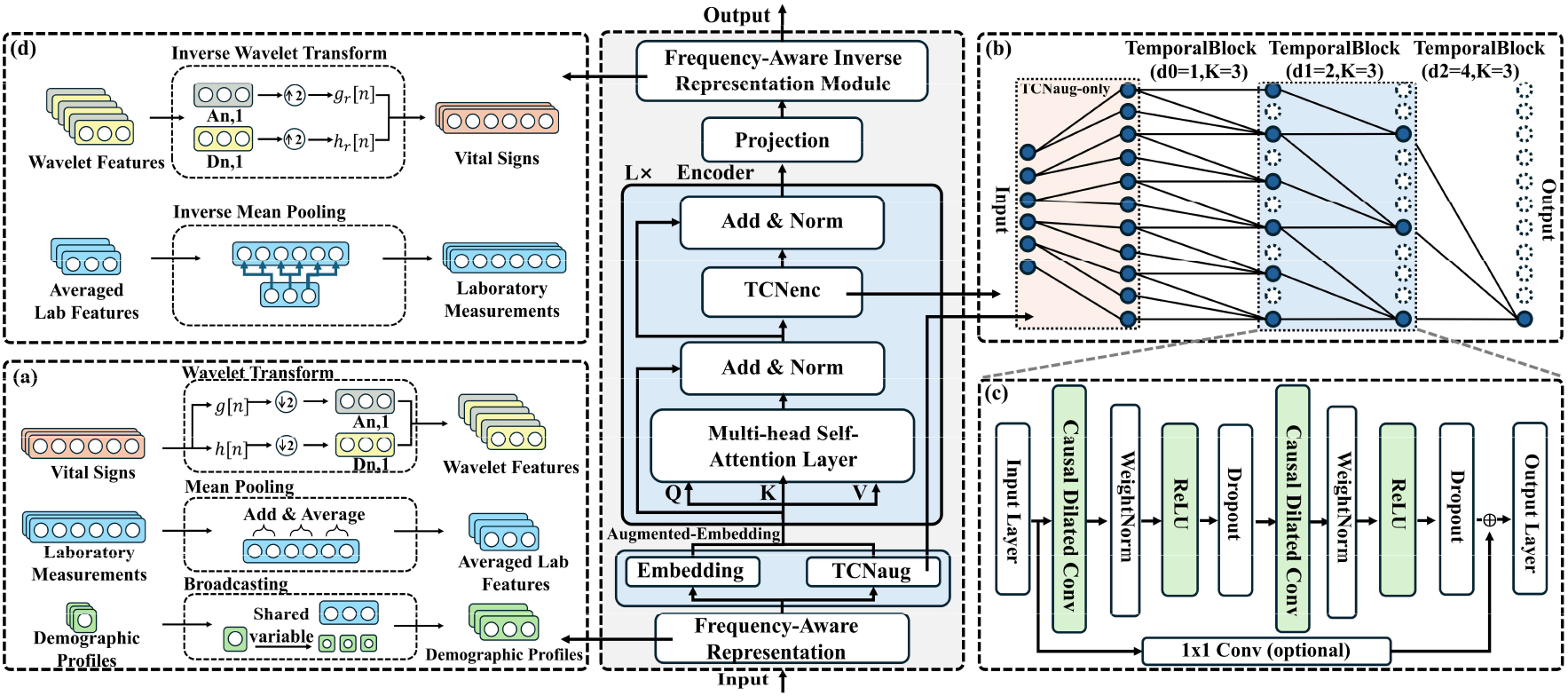
Overview and visualization of information flow in the FETT model **a** Frequency-aware representation module **b** Schematic diagram of a stack of causal dilated convolutions in TCN **c** TCN TemporalBlock **d** Frequency-aware Inverse representation module

FETT is built upon an iTransformer backbone. The original iTransformer architecture comprises an embedding module, an encoder module with *L* stacked encoder blocks, and a projection module [20]. In contrast, FETT introduces four key architectural modifications: (1) a frequency-aware representation module that extracts frequency-domain features from vital signs to complement time-domain cues (Fig. 1a); (2) an augmented embedding module that combines an embedding submodule and a TCN submodule to inject temporal order information prior to the attention mechanism (detailed in Fig. 1b and c); (3) a replacement of the FFN submodule with a TCN submodule inside each encoder block to enable multi-scale temporal modeling(detailed in Fig. 1b and c);(4) a frequency-aware inverse module that reconstructs frequency-domain predictions back into the time-domain to enhance clinical interpretability (Fig. 1d). These modifications aim to improve the iTransformer architecture from two perspectives. First, the Frequency-Aware Representation and Frequency-Aware Inverse modules function as a complementary pair, which extract frequency-domain information, incorporate it into the feature learning process, and then merge the frequency-domain predictions back into clinically interpretable time-domain outputs. Second, the introduction of two TCN submodules strengthens temporal modeling by explicitly encoding sequential order and multi-scale dependencies that standard attention mechanisms may overlook.

At a high level, given historical physiological signals *X* as input, FETT operates through the following steps.

1. Frequency-aware representation. A level-1 DWT with the db4 wavelet is applied to each vital sign signal to obtain level-1 approximation and detail coefficients. Db4 was selected following empirical comparisons among candidate wavelets, which indicated the best balance between forecast accuracy and computational cost for these tasks. Laboratory measurements are temporally aligned using mean pooling. Static demographic profiles are broadcast across all time steps, providing uniform contextual information throughout the temporal sequence. These steps produce a multi-feature time-series data *X*_*fre*_.

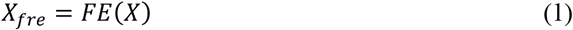

where *FE* is the frequency-domain enhancement function.
2. Augmented-embedding module. For each variable in *X*_*fre*_, the module forms embeddings through two parallel branches: a standard linear embedding branch, and a temporal branch that applies *TCN*_*aug*_ along the temporal axis. The embedding process can be defined as:

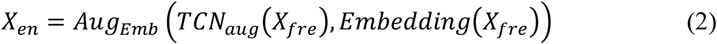

where *Aug*_*Emb*_ is the embedding augmentation function, *X*_*en*_ is the resulting embedding.
3. Encoder stack. The encoder consists of *L* stacked blocks through which data flow sequentially. The first block takes *X*^(0)^ = *X*_*en*_ as input. In each block, the multi-head self-attention layer operates on the input to model cross-variable correlations within the multivariate time-series. The output of the multi-head self-attention layer is added to the original input using residual connection to prevent vanishing gradient, followed by normalization layer to obtain a temporary representation 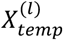:

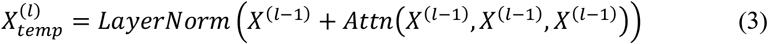

where “+” indicates a residual connection. This process is repeated in each subsequent block. *Attn* indicates the multi-head self-attention layer function. *LayerNorm* indicates the normalization layer function.

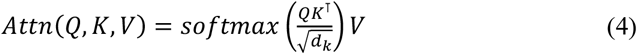

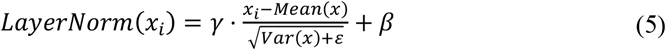

where *Mean*(*x*) indicates the arithmetic mean along the vector dimension for the current sample, *Var*(*x*) indicates the variance of the same sample over that dimension. Subsequently, 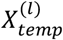 is fed into *TCN*_*enc*_ layer to capture long-range temporal dependencies and sequential patterns. The output of the *TCN*_*enc*_ layer is added to 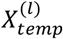 using residual connection, followed by normalization layer to yield the block output:

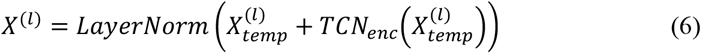

This process continues until the last block, where the final encoded representation is obtained as 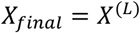.
4. Projection and inverse reconstruction. The linear projection applies a fully connected layer to *X*_*final*_, producing a multi-feature forecast 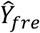 that incorporates frequency-domain features of vital signs. The projection process can be defined as:

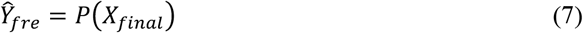

where *P* indicates the projection function. Finally, the frequency-aware representation module applies level-1 inverse DWT (db4) to vital sign components and inverse pooling to laboratory components, thereby reconstructing the time-domain trajectories:

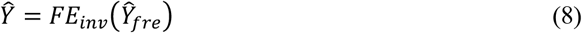

where *FE*_*inv*_ indicates the inverse frequency-domain enhancement function.

### 3.3 The frequency-aware representation module

Physiological signals, especially vital signs, contain abundant frequency-domain cues indicative of periodicity and spectral patterns. However, many models neglect these frequency patterns, limiting their ability to model periodicities and detect frequency-specific anomalies. To address this limitation, a frequency-aware representation has been introduced to extract frequency-domain features and augment the original sequence. Within this module, DWT is the core of the frequency-domain feature extraction process. Specifically, a level-1 DWT with db4 wavelets decomposes each vital sign signal *S*_*n*_ with length *T* into the approximation coefficients *A*_*n*,1_ and detail coefficients *D*_*n*,1_, using db4 low-pass filter *g* and high-pass filter *h*:

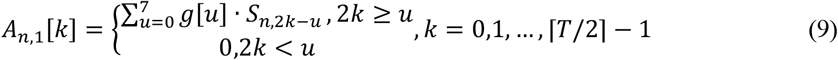

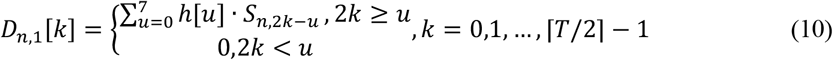

which is equivalent to convolution followed by stride-2 downsampling, and zero-padding is applied when 2*k* < *u*. As shown in Fig. 2, the low-frequency approximation coefficients 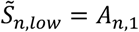 and high-frequency detail coefficients 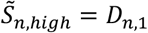 have length ⌈*T*/2⌉, because the transform performs downsampling that halves the original sequence length.

**Fig. 2.**
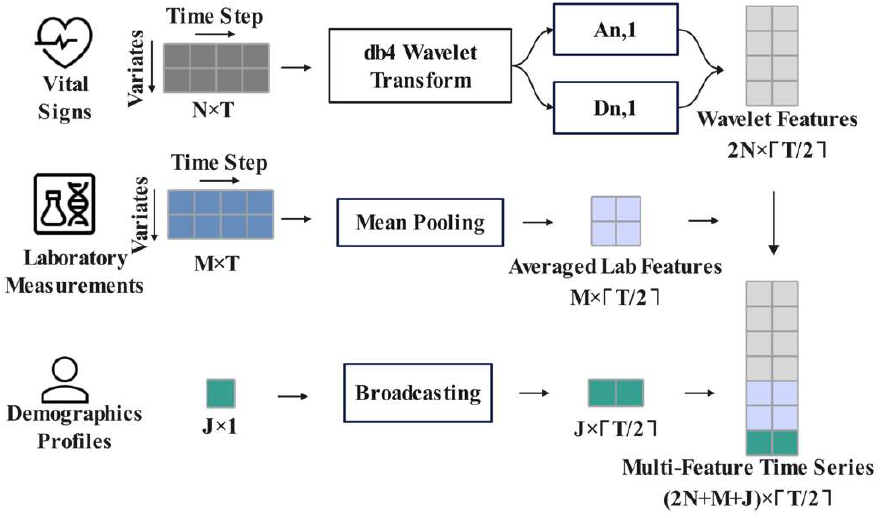
Illustration of the frequency-aware representation module

To align with the frequency-domain features of vital signs, mean pooling is applied to each laboratory measurement *L*_*m*_. Mean pooling effectively reduces the temporal resolution while retaining the key trend information. The pooling formula with a pooling window size of 2 is defined as:

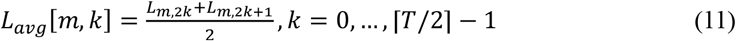

When the input time-series length *T* is odd, the signal is padded by duplicating the value of the last time step once, thereby extending its length to *T* + 1. Demographic profiles remain time-invariant; each profile is broadcast along the temporal axis to align with the sequence length, obtaining 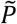.

Lastly, the time-frequency domain data of vital signs 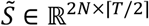, averaged laboratory measurements *L*_*avg*_ ∈ ℝ^*M*×⌈*T*/2⌉^, and demographic profiles 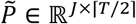 are concatenated along the variable dimension, forming the multi-feature time-series data *X*_*fre*_ ∈ ℝ^2*N*+*M*+*j*×⌈*T*/2⌉^.

### 3.4 The augmented-embedding module

The iTransformer adopts a variable-as-token formulation, in which each variable is treated as an individual token and temporal aggregation is performed via linear projection. Whereas this design enables cross-variable interactions, it lacks explicit modeling of temporal order. To address this limitation, an augmented embedding module with two parallel branches is introduced: a linear embedding submodule *Embedding* and a temporal convolutional submodule *TCN*_*aug*_. The

*TCN*_*aug*_ submodule’s core is a TCN composed of multiple TemporalBlocks (Fig. 1b). In *TCN*_*aug*_, upsampling by 2 is first applied to expand the temporal dimension of the input sequence to the embedding dimension length, as shown in the *TCN*_*aug*_-only part in Fig. 1b. Subsequently, each TemporalBlock applies two weight-normalized causal dilated convolutions (CDC), with a ReLU activation and Dropout applied after each convolution (Fig. 1c). CDC ensures the capture of long-range temporal dependencies without violating causal order. Specifically, introducing a dilation factor expands the receptive field, and the CDC at time *t* can be formulated as:

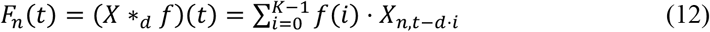

where *d* indicates the dilation factor; *K* indicates the kernel size; *t* − *d* · *i* represents the shifted index toward the past with zero-padding on the left to ensure causality.

To explicitly capture the temporal context introduced by dilation, in the *y*-th TemporalBlock (kernel size *K*, dilation factor *d*_*y*_), the (*y* − 1)-th layer output *h*^(y-1)^ is used to stack the past K samples to construct the context vector *β*:

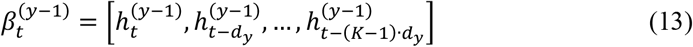

Let 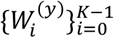 denote the per-lag convolution kernels. Concatenating the kernels along the lag dimension gives 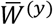. Accordingly, the CDC can be formulated as a single linear mapping followed by a nonlinear activation:

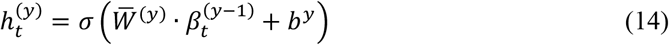

where *σ* is the activation function (ReLU), and *b*^*y*^ is the bias vector for the *y*-th TemporalBlock. The *TCN*_*aug*_ submodule and the embedding submodule jointly embed the multi-feature time-series data *X*_*fre*_ into a high dimensional feature space. This process is expressed as:

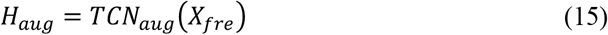

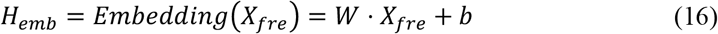

where *W* is the learnable weight matrix that expands the dimensionality, and *b* is the bias vector. The final embedding *X*_*en*_ is obtained by adding the scalar values of *H*_*emb*_ and *H*_*aug*_, note that both outputs are embedded along the temporal axis to ensure shape alignment before performing the element-wise summation.

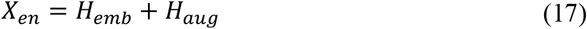

### 3.5 TCN in encoder

The augmented embedding *X*_*en*_ is fed into a stack of *L* encoder blocks. Each encoder block contains a multi-head self-attention layer and a *TCN*_*enc*_ layer. Both layers are equipped with residual connections followed by Layer Normalization for stabilizing training and improving convergence.

Each encoder block first applies multi-head self-attention across variables, allowing information to be exchanged among variables within the same processing stage and enabling the framework to capture global inter-variable dependencies. Second, unlike the design of the iTransformer, the position-wise FFN is replaced with a *TCN*_*enc*_ that operates along each token’s feature axis (Fig. 1b). Because this axis encodes temporal order within a higher-dimensional space, applying CDC within *TCN*_*enc*_ along this axis enables multi-scale temporal modeling.

The multi-head self-attention of the iTransformer backbone and the dual-TCN integration establishes a complementary architectural design for multi-scale temporal learning: Self-attention captures global dependencies among variables but lacks an inherent inductive bias toward temporal order. Placed before the encoder, *TCN*_*aug*_ operates along the true time axis and injects explicit ordinal semantics into each variable token during time-to-vector embedding. Inside the encoder, *TCN*_*enc*_ operates along the feature axis, refining multi-scale temporal patterns on top of the attention mixture. Together, these components create a synergistic flow of information:

*TCN*_*aug*_ (ordinal semantics) → Multi-head Self-attention (global interactions) → *TCN*_*enc*_ (multi-scale calibration).

### 3.6 Frequency-aware inverse representation module

To complete the frequency-aware pathway, the predicted frequency-domain components (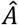 and 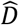) are merged back to the time-domain forecasts by applying a level-1 inverse DWT using the db4 wavelet (Fig. 1d). Specifically, the approximation coefficients and detail coefficients are up-sampled by a factor of two and convolved with the db4 low-pass reconstruction filter *g*_r_ and high-pass reconstruction filter *h*_*r*_, respectively. The resulting signals are then summed to obtain the reconstructed sequence. When the prediction length *E* is odd, the projection module maps the sequence to a length of ⌈*E*/2⌉; after the inverse transform, the sequence length becomes *E* + 1, and only the first *E* time steps are retained. For a length-8 db4 filters, when the shifted index falls outside of the valid range (i.e., *t* − 2*k* < 0 or *t* − 2*k* > 7), the corresponding entries of *h*_*r*_ and *g*_*r*_ are zero-padded. The reconstruction is computed as:

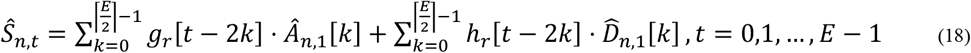

Where 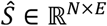. For predicted laboratory measurements 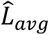, an inverse mean-pooling step is applied to restore the aggregated values to prediction length. Each pooled value is uniformly redistributed across its corresponding time window.

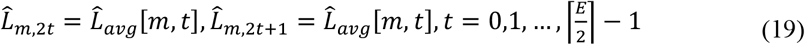

Where 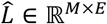. Finally, the outputs from the frequency-aware inverse representation module are concatenated along the variable dimension to yield the final time-domain predictions 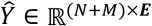.

## 4. Experiments

This section presents the evaluation of the proposed FETT framework. The evaluation is conducted on time-series forecasting tasks using electronic health records of sepsis patients from the publicly available MIMIC-IV dataset, where a framework-based model is trained, calibrated, and validated end-to-end. Sepsis is a life-threatening syndrome with dysregulated host response and organ dysfunction [34]. Its high prevalence in the ICU and rapid deterioration make it a representative target for early detection and risk forecasting.

### 4.1 Data and pre-processing

The Medical Information Mart for Intensive Care IV (MIMIC-IV) database is a large, publicly available dataset that contains de-identified hospital records from approximately 300,000 patients between 2008 and 2019. Approximately 66,000 patients were admitted to intensive care units (ICUs) [35]. Each MIMIC-IV record includes demographic profiles, vital sign measurements, laboratory test results, treatment procedures, medications, clinical notes, imaging reports, and mortality.

In the dataset, 27,207 health records of adult patients (≥18 years) who met the Sepsis-3 criteria [34] and with at least one ICU stay were included, and only the first ICU stay for each patient was used. The following preprocessing steps were performed to prepare the data for subsequent experiments:

1. To ensure data integrity and avoid bias, variables with insufficient availability were excluded. Specifically, first, vital sign variables with more than 20% missing values were removed from the variable list. Second, given the intermittent and inherently sparse nature of laboratory testing, lab variables with insufficient availability were removed. Missing segments were imputed using last observation carried forward within the observation window to avoid information leakage.
2. Signals were resampled to an hourly grid. For highly variable indicators (e.g., heart rate, respiratory rate), an hourly trimmed mean was computed by excluding the within-hour minimum and maximum; for more stable variables, a simple hourly mean was used.
3. The first 72 hours of each ICU stay were extracted, and episodes shorter than 72 hours were excluded.
4. For eligible records, a block-wise sliding-window strategy was employed. The look-back window had a length of *L*, the forecast window had a length of *H*, and the stride was set to *s* = *L* + *H*. Both the look-back and forecast windows were non-overlapping, ensuring that consecutive samples were temporally independent and preventing any potential temporal leakage.
5. Each variable was standardized using the training set statistics:

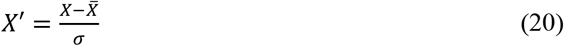

where 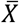 indicates the mean of the data, and *σ* indicates the standard deviation.

After preprocessing, 23,504 sepsis patient records were retained, each containing 26 standardized variables (As listed in Table 2) collected during the first 72 hours of ICU admission. Data were split into training, validation, and test sets in an 8:1:1 ratio. The records were randomly assigned to each split based on unique hospital admission identifiers, ensuring episode-level independence across the datasets.

**Table 2.**
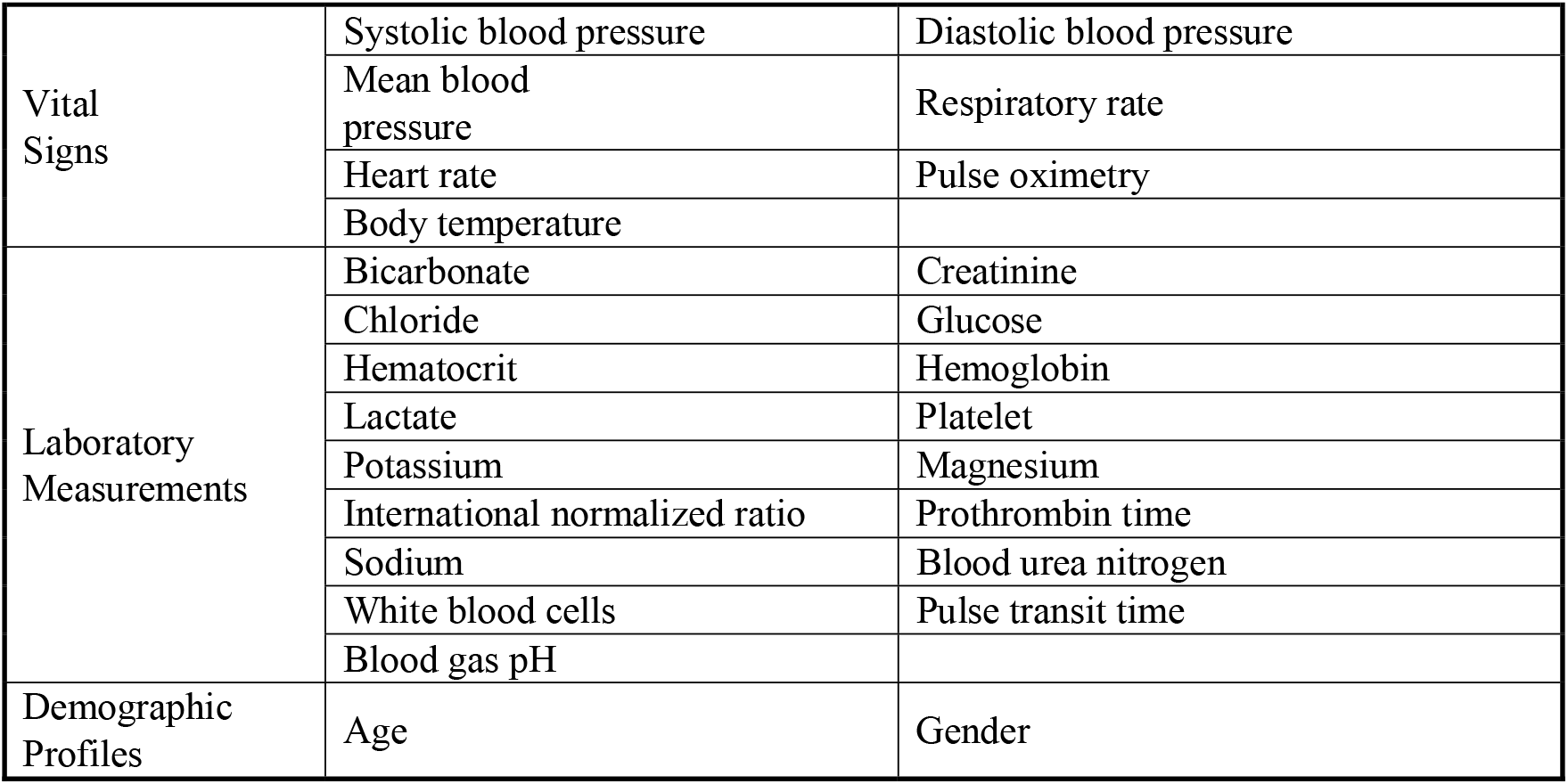
Categorization and summary of clinical variables included in the MIMIC-IV sepsis cohort.

### 4.2 Implementation

The experiment was conducted using PyTorch. Each input is embedded into a 128-dimensional vector. The encoder comprises two Transformer encoder blocks (*L* = 2), with each using four self-attention heads.

Both *TCN*_*aug*_ and *TCN*_*enc*_ in the framework consist of 4 layers of TemporalBlocks with kernel size *K* = 5. The dilation factors increase exponentially (e.g., 1, 2, 4, 8).

Training proceeded for up to 50 epochs with early stopping (patience = 5) based on validation loss to prevent overfitting. Training has been conducted with AdamW (learning rate 1e-3, weight decay 1e-5), a batch size of 128, mean squared error (MSE) as the loss for continuous-value forecasting, and a dropout rate of 0.1. Hyperparameters were chosen via grid search with cross-validation on the training set. Unless otherwise noted, all baselines used the same configuration, and the same train/validation/test splits as our framework. Experiments were conducted on an NVIDIA GeForce RTX 4080 GPU (16 GB) using PyTorch 2.4.0 and CUDA 12.7.

### 4.3 Baseline methods and evaluation metrics

The methods compared are summarized as follows. RNN [15], GRU [36], LSTM [37], Transformer (self-attention) [38], Informer (sparse Transformer for long sequences) [21], TimesNet (2D variation modeling for time series) [33], iTransformer (inverted attention design) [20], TimeXer (Transformer with exogenous variables) [39], and FEDformer (frequency-domain enhanced Transformer) [22].

In comparative experiments, to evaluate the accuracy of FETT in predicting sepsis progression across different time spans, two prediction strategies were employed: using the first 12 hours of data to predict the next 12 hours and using the first 24 hours to predict the next 24 hours. The evaluation metrics used in this experiment are Mean Squared Error (MSE) and Mean Absolute Error (MAE). MSE is defined as the average of the squared differences between the predicted and actual values, measuring the overall magnitude of prediction errors. It places greater emphasis on large deviations, making it more sensitive to outliers.

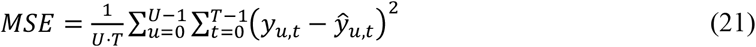

MAE is defined as the average of the absolute differences between the predicted and actual values, providing a straightforward measure of the average prediction deviation. It provides an intuitive measure of average deviation while being more robust to outliers.

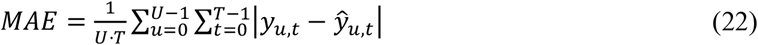

where *y*_*u,t*_ indicates the real value of the variable *u* at time *t*; ŷ_*u,t*_ indicates the value of the variable *u* predicted at time *t*; *U* = *N* + *M* is the total number of variables.

Together, these two metrics provide a comprehensive assessment of framework performance by jointly capturing both the overall prediction accuracy and the sensitivity to large deviations.

## 5. Results and discussion

### 5.1 Comparative results on sepsis forecasting tasks

Table 3 presents a detailed performance comparison of all models on the sepsis dataset. The best results are highlighted in bold, and the second-best results are underlined. The downward arrows indicate the relative performance improvement achieved by the proposed method compared with each baseline.

**Table 3.** Comparative performance evaluation of FETT and baseline models on sepsis progression forecasting tasks.

| Methods | 12 Hours |  | 24 Hours |  |
| --- | --- | --- | --- | --- |
|  | MSE | MAE | MSE | MAE |
| RNN | 0.3465 | 0.3593 | 0.4283 | 0.4319 |
| LSTM | 0.3401 | 0.3547 | 0.4558 | 0.4470 |
| GRU | 0.3236 | 0.3484 | 0.3857 | 0.4165 |
| Transformer | 0.2611 | 0.2828 | 0.3152 | 0.3374 |
| Informer | 0.2478 | 0.2692 | 0.3115 | 0.3412 |
| TimeXer | 0.2628 | 0.2580 | 0.3338 | 0.3289 |
| TimesNet | 0.2763 | 0.2687 | 0.3501 | 0.3367 |
| iTransformer | 0.2511 | 0.2587 | 0.2959 | 0.3121 |
| FEDformer | 0.2663 | 0.2787 | 0.3102 | 0.3266 |
| Ours | **0.2172** | **0.2408** | **0.2343** | **0.2698** |
| vs. iTransformer | 13.50%↓ | 6.91%↓ | 20.81%↓ | 13.55%↓ |
| vs. Best Baseline | 12.34%↓ | 6.67%↓ | 20.81%↓ | 13.55%↓ |

In the 12-hour task, FETT achieves an MSE of 0.2172 and an MAE of 0.2408. These results represent error reductions of 12.35% and 6.67% relative to the strongest baselines—Informer (MSE = 0.2478) and TimeXer (MAE = 0.2580), respectively. In the 24-hour task, FETT achieves an MSE of 0.2343 and an MAE of 0.2698, improving over the strongest baseline iTransformer (MSE 0.2959, MAE 0.3121) by 20.82% and 13.55%. The performance gap widens from 12.35%/6.67% (12 h) to 20.82%/13.55% (24 h), indicating better long-horizon modeling. Recurrent baselines (RNN, LSTM, and GRU) exhibit substantially higher errors—for example, RNN records an MSE of 0.4283 in the 24-hour task. Whereas attention-based baselines (Transformer and Informer) perform competitively in the 12-hour task (e.g., Informer achieves an MSE of 0.2478), their errors increase more substantially in the 24-hour task. Overall, FETT attains the lowest errors across both horizons. Reported results represent means over multiple runs.

### 5.2 Ablation studies

Ablation studies were conducted to evaluate the contributions of the three key innovative components of FETT: (1) two TCN components (*TCN*_*aug*_ and *TCN*_*enc*_), (2) the frequency-aware representation module, (3) the wavelet family and decomposition level used inside the frequency-aware representation module.

First, to evaluate the impact of *TCN*_*aug*_ and *TCN*_*enc*_ on the framework performance, the following ablation experiment schemes were designed:

1. FETT w/o TCN: Both *TCN*_*aug*_ and *TCN*_*enc*_ are removed, reducing the framework to the attention-based backbone combined with the frequency-aware representation module.
2. FETT w/o *TCN*_*enc*_: Only the *TCN*_*aug*_ component is retained.
3. FETT w/o *TCN*_*aug*_: Only the *TCN*_*enc*_ component is retained.

The test results of the ablation experiments are shown in Table 4. The results show that FETT w/o TCN performs well in 12-hour task but shows a significant increase in MSE and MAE for 24-hour tasks, underscoring iTransformer’s limitations in capturing extended trends.

**Table 4.** Results of ablation studies quantifying the contributions of the dual-TCN design and frequency-aware representation module.

| iTransformer | $TCN_{aug}$ | $TCN_{enc}$ | Frequency-aware representation | 12 Hours | | 24 Hours | |
| --- | --- | --- | --- | --- | --- | --- | --- |
|  |  |  |  | MSE | MAE | MSE | MAE |
| ✓ |  |  | ✓ | 0.2343 | 0.2420 | 0.2845 | 0.3019 |
| ✓ | ✓ |  | ✓ | 0.2238 | 0.2420 | 0.2688 | 0.2854 |
| ✓ |  | ✓ | ✓ | 0.2269 | 0.2553 | 0.2540 | 0.2951 |
| ✓ | ✓ | ✓ |  | 0.2346 | 0.2542 | 0.2589 | 0.2933 |
| ✓ | ✓ | ✓ | ✓ | 0.2172 | 0.2408 | 0.2343 | 0.2698 |

FETT w/o *TCN*_enc_ reduces MSE and MAE to 0.2238 and 0.2420 for the 12-hour task. For the 24-hour task, MSE and MAE are reduced to 0.2688 and 0.2854. These results highlight the critical role of *TCN*_*aug*_ in improving accuracy across both horizons.

FETT w/o *TCN*_*aug*_ also enhances performance, with MSE and MAE decreasing to 0.2269 and 0.2553 for 12-hour task. For the 24-hour task, MSE and MAE decrease to 0.2540 and 0.2951. Combining both *TCN*_aug_ and *TCN*_enc_ yields the best performance, reducing MSE and MAE to 0.2172 and 0.2408 in 12-hour task. For the 24-hour task, MSE and MAE are reduced to 0.2343 and 0.2698. This demonstrates that whereas iTransformer provides the foundational time-series modeling, the TCN components enhance the capture of global temporal dependencies, working synergistically to improve FETT’s overall effectiveness.

Next, to evaluate the impact of the frequency-aware representation module, FETT was tested without it. As shown in Table 4. Without the module, the framework performs well for 12-hour task but experiences substantial MSE and MAE spikes in 24-hour task. Adding the module reduces errors in both 12- and 24-hour tasks, with particularly strong reductions of 9.50% in MSE and 8.01% in MAE for 24-hour task.

Finally, wavelet choices were compared. As shown in Table 5, db4 outperformed Haar, db2, and db6, yielding the lowest errors across both horizons. For db4, decomposition levels 1, 2, and 3 were evaluated. Since only one wavelet transform is performed, level-1 requires less computational effort compared to deeper levels, resulting in shorter training time. Level-1 achieves the best MSE/MAE with minimal training time.

**Table 5.**
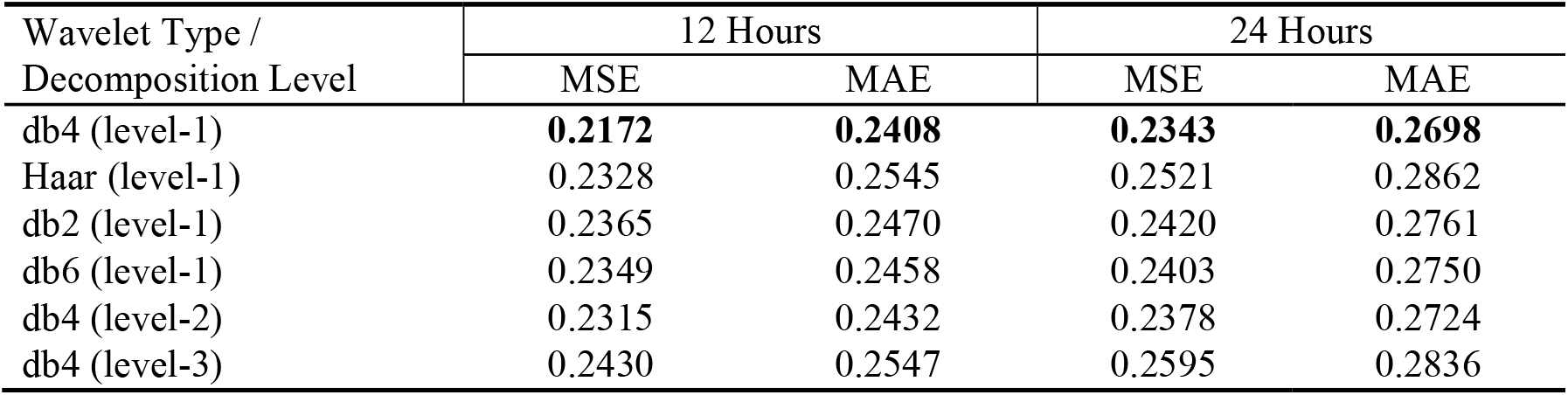
Performance comparison of different wavelet families and decomposition levels within the frequency-domain stream.

### 5.3 Prediction performance on different time steps

In the 24-hour prediction task, prediction errors were evaluated at each time step to analyze their temporal trend. As shown in Fig. 3, the analysis focuses on the first 12 time steps to capture the initial rapid rise in error and its subsequent stabilization phase. Beyond the 12th step, errors remain relatively stable, contributing limited additional insight. Therefore, only the first 12 time steps are presented in Fig. 3.

**Fig. 3.**
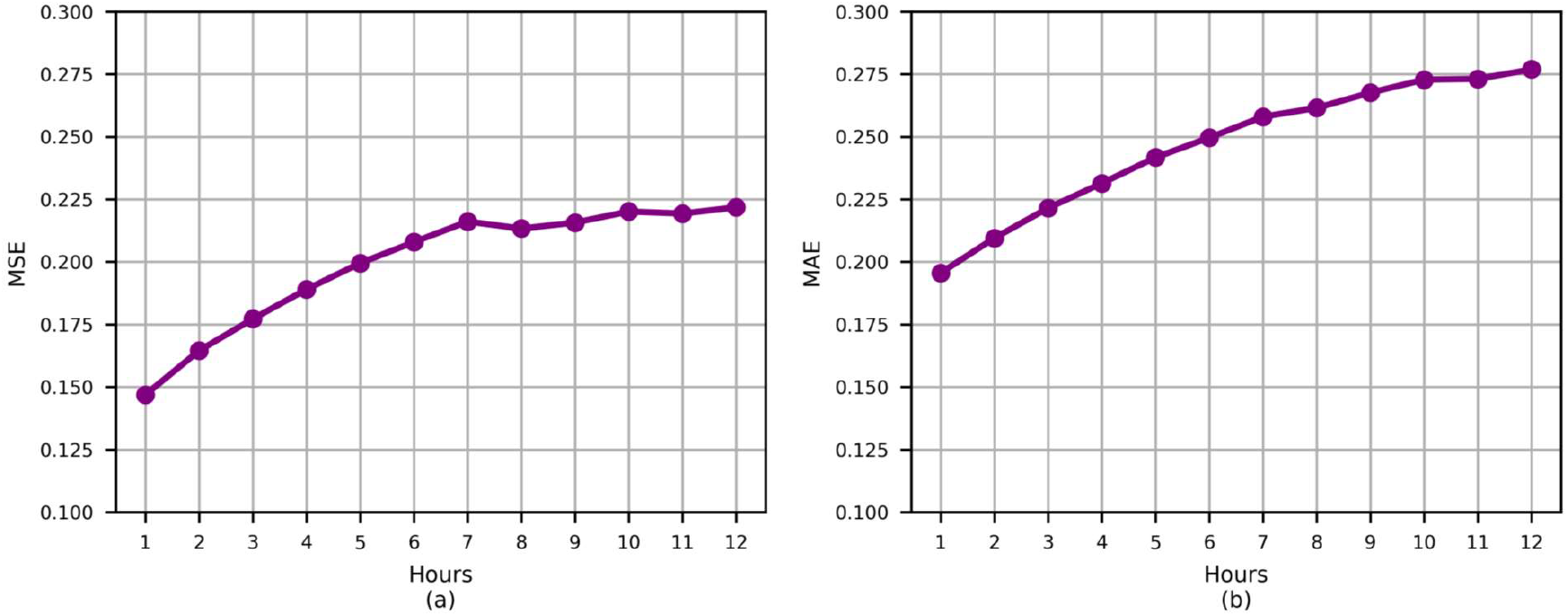
Analysis of the model’s performance across various time steps within the first 12 hours of the 24-hour sepsis dataset prediction task

For MSE (Fig. 3a), the error starts at 0.1469 for the first forecast step and rises to 0.2079 at the sixth step, representing a substantial increase of 41.52% relative to step 1. It then grows more gradually, reaching 0.2218 at the twelfth step—an additional 6.69% increase compared to step 6. This trend suggests that the framework maintains high predictive accuracy at the initial horizons, with performance degradation becoming less pronounced over longer forecasting intervals. MAE exhibits a similar trend (Fig. 3b), further corroborating the increase in prediction error with longer forecasting horizons, whereas the error stabilizes after mid-horizon. This suggests that clinicians should interpret predictions with progressively lower confidence at longer forecasting horizons, while relying more on early-horizon outputs for high-confidence decision-making.

### 5.4 Number of TemporalBlocks in TCNs

TemporalBlocks serve as the core computational units of the TCN components, determining the framework’s receptive field. In this experiment, we investigate how varying the number of TemporalBlocks affects both prediction accuracy and computational efficiency.

In the 24-hour prediction task, experimental results show that when the number of TemporalBlocks in either TCN exceeds 6, the prediction error steadily increases. Therefore, for clearer visualization and discussion, Figure 4 shows the results with no more than 6 TemporalBlocks in either TCN. Additionally, comprehensive experiments were conducted by fixing the number of TemporalBlocks in one TCN between 1 and 6 while varying it in the other over the same range. The results show that the prediction error reaches its minimum when both TCNs contain four TemporalBlocks (MSE = 0.2343, MAE = 0.2698). Accordingly, Figure 4 illustrates the MSE and MAE values obtained by varying the number of TemporalBlocks in one TCN from 1 to 6, while keeping the other fixed at 4. When the depth deviates from this configuration, performance degrades. For example, with *TCN*_enc_ fixed at four TemporalBlocks, configurations with one (MSE = 0.2481, MAE = 0.2845) or six (MSE = 0.2349, MAE = 0.2736) layers yield higher errors. Similarly, when *TCN*_*aug*_ is fixed at four TemporalBlocks, configurations with one (MSE = 0.2490, MAE = 0.2829) or six (MSE = 0.2359, MAE = 0.2743) layers also perform worse.

**Fig. 4.**
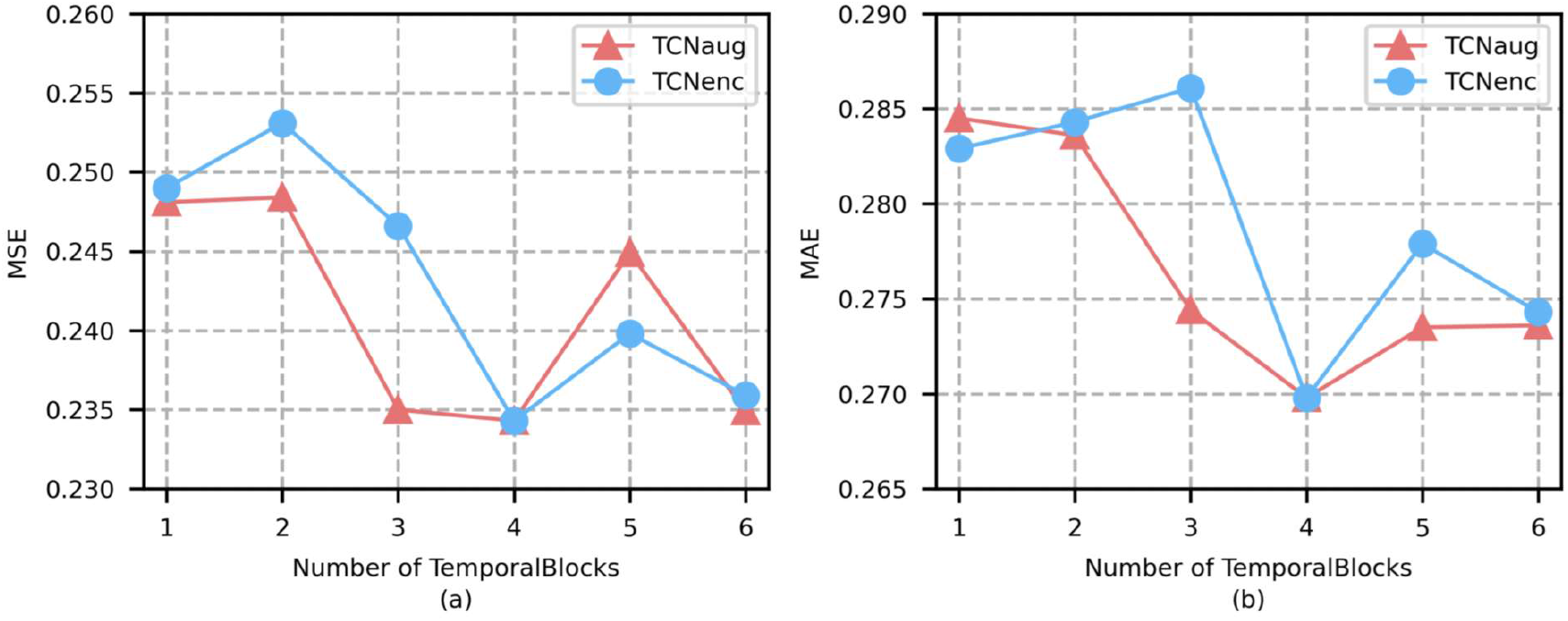
Impact of the number of TemporalBlocks on the FETT in the 24-hour prediction task of the sepsis dataset

This result suggests that there is an optimal number of TemporalBlocks in TCN. Excessive TemporalBlocks may cause overfitting, increasing computational demands without substantially improving predictive accuracy, whereas too few TemporalBlocks may not be able to capture the complex temporal dependencies in the data, leading to a decrease in the framework’s fitting capability.

## 6. Conclusion

This paper proposes an innovative multivariate time-series forecasting framework named FETT (Frequency-Enhanced TCN-Transformer), a unified framework that integrates temporal and frequency-domain of physiological signals for disease progression prediction in critically ill patients. It combines the multi-head self-attention design of iTransformer and a dual TCN structure to capture global and multi-scale temporal patterns of physiological signals. Additionally, a frequency-aware representation module decomposes the frequency characteristics of vital signs and integrates laboratory data and demographic profiles for comprehensive analysis of time- and frequency-domain information. Experiments on the MIMIC-IV dataset showed that FETT outperformed existing models in sepsis-progression prediction, with reductions in MSE and MAE for both 12-hour and 24-hour tasks. These results demonstrate its effectiveness for both short- and long-term disease-progression prediction.

The FETT framework holds significant potential for clinical applications, particularly in critical care settings such as intensive care units (ICUs) and emergency departments. It can be seamlessly integrated with real-time vital sign monitors and electronic health record (EHR) systems to analyze multivariate time-series data encompassing vital signs and laboratory measurements. By leveraging these data, FETT enables dynamic prediction of disease progression in critically ill patients, allowing clinicians to identify early signs of physiological deterioration and initiate timely interventions—often several hours before overt clinical decline. Furthermore, the predicted physiological trajectories (e.g., heart rate, respiratory rate, blood pressure) can be mapped to established early-warning systems such as NEWS or EWS, enabling continuously updated risk scores. Similarly, organ-specific trajectories allow estimation of SOFA score trends, providing a more granular and forward-looking assessment compared with traditional snapshot scoring [34, 40, 41]. This functionality assists clinicians in prioritizing patient monitoring, escalating therapy, and initiating rapid-response evaluations. Finally, FETT performs forecasting of future vital sign variations through the integration of frequency-domain reconstruction and time-domain fusion mechanisms. The predicted trajectories align closely with the periodic physiological signal fluctuations observed by clinicians, thereby providing a valuable foundation for enhancing the interpretability of the framework’s outputs.

Despite its advantages, there are still some limitations of this work. First, although FETT demonstrates strong predictive performance, its complex architecture entails substantial computational overhead during both training and inference. The increased number of parameters and intermediate activations leads to higher GPU memory consumption, which may pose challenges for deployment in resource-constrained environments. Second, the current implementation utilizes only single-modal medical time-series data, without incorporating other modalities such as clinical notes or imaging studies. This restriction may limit the framework’s capacity to capture complementary information across heterogeneous clinical data sources. To address these limitations, future work will advance along two directions: (1) streamlining the architecture through model-compression techniques (e.g., knowledge distillation and structured sparsity) to improve computational efficiency and reduce memory consumption; and (2) integrating multimodal data and applying transfer learning to enhance predictive accuracy and robustness, while prospectively calibrating framework outputs against SOFA and NEWS/EWS trajectories to ensure that FETT’s predictions remain directly interpretable and aligned with established clinical scoring systems.

## Data Availability

All data produced in the present study are available upon reasonable request to the authors

## Declarations

### Ethical approval

This study utilizes the publicly available, de-identified MIMIC-IV electronic health record dataset. All data have been de-identified in compliance with Health Insurance Portability and Accountability Act (HIPAA) standards to ensure patient privacy and confidentiality. Access was granted upon completion of the required Collaborative Institutional Training Initiative (CITI) “Data or Specimens Only Research” course and registration with PhysioNet. As the dataset is fully de-identified and does not involve human subjects, this study is exempt from institutional review board (IRB) approval. The research adheres to the ethical principles outlined in the Declaration of Helsinki and complies with relevant data use agreements.

### Data availability

Publicly available datasets were analyzed in this study. The datasets are available on the MIMIC-IV website at: https://physionet.org/content/mimiciv/2.2/

